# Exome first approach to reduce diagnostic costs and time – retrospective analysis of 111 individuals with rare neurodevelopmental disorders

**DOI:** 10.1101/2021.07.23.21258974

**Authors:** Julia Klau, Rami Abou Jamra, Maximilian Radtke, Henry Oppermann, Johannes R. Lemke, Skadi Beblo, Bernt Popp

## Abstract

This single center study aims to determine the time, diagnostic procedure and cost saving potential of early exome sequencing in a cohort of 111 individuals with genetically confirmed neurodevelopmental disorders.

We retrospectively collected data regarding diagnostic time points and procedures from the individuals’ medical histories and developed criteria for classifying diagnostic procedures in terms of requirement, followed by a cost allocation. The genetic variants of all individuals were reevaluated according to ACMG recommendations and considering the individuals’ phenotype.

Individuals who developed first symptoms of their underlying genetic disorder when Next Generation Sequencing (NGS) diagnostics were already available received a diagnosis significantly faster than individuals with first symptoms before this cutoff. The largest amount of potentially dispensable diagnostics was found in genetic, metabolic, and cranial magnetic resonance imaging examinations. Out of 407 performed genetic examinations, 296 (72.7%) were classified as potentially dispensable. The same applied to 36 (27.9%) of 129 cranial magnetic resonance imaging and 111 (31.8%) of 349 metabolic examinations. Dispensable genetic examinations accounted 302,947.07€ (90.2%) of the total 335,837.49€ in potentially savable costs in this cohort. The remaining 32,890.42€ (9.8%) are related to non-required metabolic and cranial magnetic resonance imaging diagnostics. On average, the total potentially savable costs in our study amount to €3,025.56 per individual.

Cost savings by first tier exome sequencing lie primarily in genetic, metabolic, and cMRI testing in this German cohort, underscoring the utility of performing exome sequencing at the beginning of the diagnostic pathway and the potential for saving diagnostic costs and time.

## INTRODUCTION

Neurodevelopmental disorders (NDD) and epilepsy are frequent causes for medical presentations and diagnostics.^1,2^ NDD affect approximately 3% of children worldwide while about 1 in 150 children develops epilepsy in the first 10 years of life.^3,4^ Both conditions comprise very heterogeneous etiologies that include several hundred rare genetic disorders.^5–8^

Establishing a diagnosis involves coordinated interplay between pediatricians, neurologists, geneticists and other physicians in centers for rare diseases. This complex process is often highly time consuming, requires great effort through a huge amount of diagnostic procedures and immensely burdens affected families.^9–12^ The associated diagnostic costs and risks are considerable, underscoring the scope of this “diagnostic odyssey” and the load on the health care system.^13^ The impact of a conclusive genetic diagnosis for affected individuals and families has been the subject of several studies and demonstrates the importance of developing guidance for an effective diagnostic pathway.^9,14–16^

Genetic diagnostics have traditionally been initiated at a relatively late stage in the diagnostic pathway and were often performed in a stepwise manner.^17,18^ Since its introduction, high-throughput sequencing has proven to be an effective, rapid but cost-intensive method in genetic diagnostics^19^ and has evolved from a research method to a routine diagnostic tool.^20^ Next Generation Sequencing (NGS) outperforms traditional genetic diagnostics, as it can achieve a diagnostic yield between 30% and 47% in NDD and epilepsy cohorts combined with speed-up through a single test.^18,21–24^ Thus, the question remains^25^ whether first-line initiation of exome sequencing (ES) could save costs and time, reduce the risks associated with thereby obsolete extensive or even invasive diagnostic procedures, and allow families to make earlier reproductive decisions.

To address these questions, we designed a retrospective single-center study focusing on diagnostic procedures and time durations in 111 individuals with NDD and/or epilepsy who obtained a molecular diagnosis through NGS methods. We wanted to determine the extent to which NGS diagnostics influenced the duration of the diagnostic odyssey and to assess the number of diagnostic interventions that might have not been required retrospectively if ES had been implemented earlier in this cohort.

## MATERIALS AND METHODS

### Ethics Approval

The project was approved by the ethic committee of the University of Leipzig, Germany (224/16-ek and 402/16-ek) and was conducted in concordance to the declaration of Helsinki. Individuals or their parents or legal guardians consented to genetic testing.

### Study design and patient selection

We retrospectively selected 111 individuals with pediatric-onset NDD and/ or epilepsy from a diagnostic cohort of 2128 cases based on a series of filtering and quality control steps. All individuals had received a molecular diagnosis in this regard using NGS methods at the Institute of Human Genetics at Leipzig University Medical Center (UKL), a tertiary care centre in Germany. The participants received their diagnoses between 2017-04-04 and 2020-08-03. Further details on inclusion and exclusion criteria are provided in Figure 1A. These criteria are intended to ensure that physician letters were available for all individuals to collect data on diagnostic procedures as well as time points of interest. Therefore, we excluded individuals who received no clinical assessments at UKL and those born before the year 2000. To assure that only individuals with sufficiently clarified genetic diagnoses were included, we excluded cases where only variants of uncertain significance (VUS) had been reported in the initial diagnostic report. This resulted in a list of 112 individuals for which we collected clinical data. After systematic variant reevaluation in the course of this project, we excluded one individual where the previously reported variant does not sufficiently explain the severity of the case (Ind076; for further details see Supplemental File S3).

**Figure 1.**
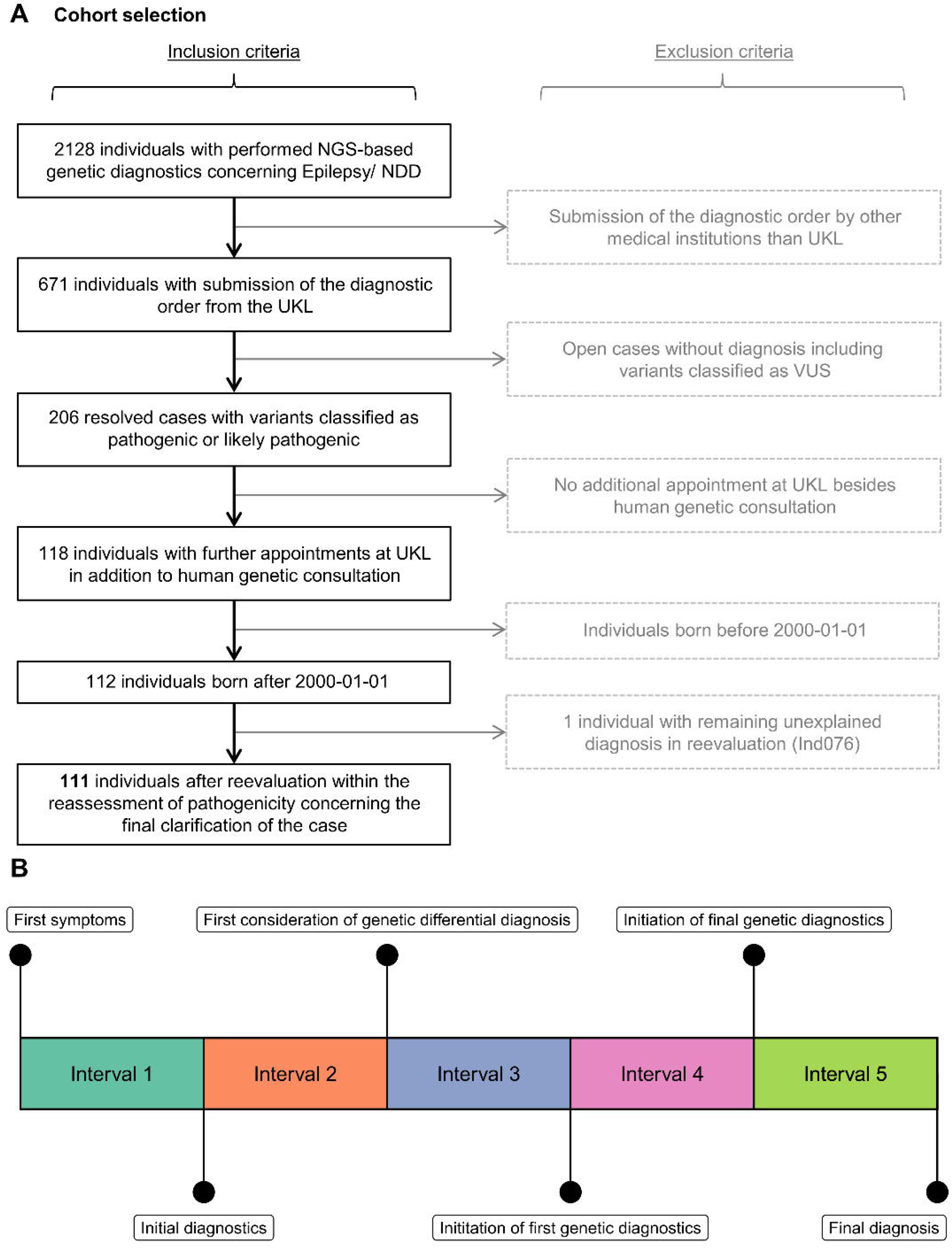
Flowchart of participant recruitment and timeline. **(A)** Workflow of the approach for selecting participating individuals for this study using the internal patient information system of the Institute of Human Genetics at the UKL. Further information on the excluded individual (Ind076) is provided in supplementary files. **(B)** Schematic timeline showing the time points of the diagnostic trajectory researched from the patient’s medical history and the resulting time intervals.

### Clinical data collection and classification

The internal patient information systems of the UKL were used to curate genetic and clinical information from the individuals’ medical history. We collected data on all diagnostic measures i.e. genetic analyses, cranial magnetic resonance imaging (cMRI), metabolic diagnostics (defined as analyses of parameters in a blood, urine or liquor sample, such as organic acids, amino acid profiles, acylcarnitines, etc.), other laboratory diagnostics, lumbar punctures, other medical imaging than cMRI (including sonographic, endoscopic, X-ray and computer tomography examinations), electrocardiograms (ECG), electroencephalograms (EEG), electrophysiology, function tests (including spirometry, polysomnography and cardiorespirogram examinations) and conciliar examinations. Metabolic parameters examined in cerebrospinal fluid samples were assigned to metabolic diagnostics. The lumbar puncture itself and the associated laboratory tests concerning, for example, inflammatory parameters are included in the lumbar puncture category. For details on the specific types of the respective diagnostic procedures, see Supplemental File S2^26^ (sheet “costs”). We also included hospitalisations and counted the amount of inpatient overnight stays. For laboratory tests other than metabolic diagnostics, we only recorded whether they were performed at least once due to the amount of single data points with neglectable overall costs. We concentrated on the diagnostics performed at UKL excluding additional examinations potentially performed at other medical institutions. This approach is in-line with previous studies^27^ and served to ensure a uniform approach because, especially in the case of individuals with a longer medical history at other hospitals, our access to physician letters was restricted. However, we included data on cMRI and genetic examinations performed in other medical institutions, because these information were consistently transmitted to the UKL. Our data collection involved all procedures performed from birth to the report of the final molecular diagnosis. We classified each diagnostic entry according to its requirement considering the individual medical history and the exact circumstances of each specific situation based on assessment by pediatricians experienced with rare diseases. Many diagnostic interventions were regarded as indispensable, especially for critically ill individuals or when warning signs like a reduced general condition, developmental regression, and new onset seizures were present. Our classification system was divided into three parts and contains the categories “not required”, “required” and “ambiguous”. All genetic investigations were classified as dispensable, except for the final NGS-based investigations leading to the diagnosis and their subsequent validation by Sanger, MLPA, and/ or q/RT-PCR. The curated classification criteria for all procedures are provided in Supplemental File S2^26^ (sheet “criteria”).

### Diagnostic time data collection

We recorded diagnostic time points for each individual based on the structured diagnostic pathway as depicted schematically in Figure 1B. Data for the following time points were collected: occurrence of first symptoms associated with the genetic phenotype (t1), initial diagnostics related to the underlying genetic disease (t2), first consideration of a genetic differential diagnosis (t3), initiation of first genetic diagnostics (t4), enrollment of the NGS-based examination that lead to the final diagnosis (t5) and the corresponding diagnosis report date (t6).

### Costs

Subsequent to the clinical data collection, we determined associated diagnostic costs using a retrospective bottom-up approach. This refers to our concept of inferring the total cost to the health care system from determining the cost of individual procedures. We focused on direct costs incurred solely due to diagnostic procedures. Other direct medical costs as well as indirect costs were not considered. The cost calculation is based on the “Gebührenordnung für Ärzte” (GOÄ), which determines the billing of medical costs in the outpatient sector in Germany. Even though many diagnostic measures performed on the individuals in our cohort occurred in a hospital setting, we nevertheless calculated them on the basis of the GOÄ. In Germany, costs incurred during hospitalizations are billed as flat-rate payments via Diagnosis Related Groups (DRGs), which does not allow conclusions to be drawn about particular diagnostic costs.^3^ Therefore, we also did not include the costs of hospitalization in our cost calculations to avoid duplications. We focused the cost analysis on the three categories with the largest amounts of non-required procedures, which were genetic diagnostics, cMRI, and metabolic diagnostic costs. Our estimated costs for gene panel sequencing, trio ES and single ES were fixed to 3461.45€ per examination based on the GOÄ cost system in September 2020 including accounting of the nucleic acids isolation from the blood sample, sequencing and scientific evaluation and report. This cost level is consistent with estimated costs of NGS-based diagnostics in other studies. Schwarze et al.^28^ report a range of costs for ES between 555$ and 5,169$ (inflated value in €: 507,79€ - 4729,30€); Vrijenhoek et al.^22^ state 3,600€ (inflated value: 3,693.27€) for ES consistent with our costs. We inflated the costs for comparative purposes using a web inflation tool (https://www.inflationtool.com) and converted them subsequently to Euro using corresponding daily exchange rates from Bloomberg L.P. on 2021-05-06 (for conversion and inflation calculation of costs see Supplementary Table S1). A specific diagnostic costs summary is available in Supplementary File S2^26^ (sheet “costs”).

### Genetic Analyses and Variants Reevaluation

All individuals were diagnosed by NGS-based examinations such as gene panel, single ES or trio ES. To increase sample size, we included individuals diagnosed by gene panel examinations in our cohort because we assumed the identified variants were definitively diagnostic and would have been detected by exome sequencing as well.^23^ Only those individuals with variants classified as pathogenic or likely pathogenic and considered to fully cause the phenotype were selected for this study. We performed a reevaluation of all detected variants according to the updated guidelines provided by the American College of Medical Genetics and Genomics (ACMG)^29^ to standardize and update all variant pathogenicity assessments. For consistent variant nomenclature and standardized evaluation, web resources like ClinVar, Varsome, Decipher, Mutalyzer, gnomAD, ClinGen Pathogenicity Calculator and AutoPVS1, were used. We ensured that all variants reported here were submitted to ClinVar (see Supplementary File S3^26^ for ClinVar IDs).

### Statistical analysis and Data Plotting

We analysed and graphically processed the data compiled in Excel (Microsoft Corporation, Redmond, Washington, USA) using R language (Version 4.0.5) from within RStudio (Version 1.4). Because of its infrequent application, the diagnostic entries that were marked in the “ambiguous” category were assigned to the “required” category for our statistical analysis following a conservative evaluation approach. In order to make comparisons of diagnostic times intervals we subdivided the cohort. This division was based on the onset time of first symptoms concerning the genetic phenotype before and after the introduction of NGS-based diagnostics. This cut-off was set to April 1, 2016, the date since when NGS based diagnostics where broadly applied at our Institute of Human Genetics in Leipzig. We used two sided Wilcoxon signed-rank test as implemented in R to calculate p-values for differences between groups.

## RESULTS

### Cohort demographics

Of the 111 individuals included, 49 (44.1 %) were female and 62 (55.8 %) were male (ratio: 1:1.26). Due to the inclusion criteria described earlier (Figure 1A), all individuals enrolled were under 20 years of age at the time of diagnosis. Details of the sex-specific age distribution are provided in Figure 2A. Initial clinical diagnosis was NDD in 69 (62.1%) of individuals, epilepsy in 10 (9%), and a combined occurrence of both in 32 (28.8%) (Figure 2B). In 10 individuals with isolated epilepsy the seizure onsets and genetic diagnoses occurred in a very young age, or no medical records were available for the time after genetic diagnosis. We examined further medical letters for possible later onsets of NDD, but no evidence for it was found. Thus, it cannot be excluded that NDD could still develop in these individuals later in life. The onset of the first symptoms associated with the underlying genetic disease developed at a median age of 6.2 months (range: 0 – 156.6; standard deviation (SD): 20.5) in the individuals. Out of 111 individuals 22 individuals (19.8%) were already symptomatic regarding the molecular diagnosis on their day of birth and 50 individuals (45%) showed symptoms in their first year of life; another 39 individuals (35.1%) became symptomatic in their second year of life or later. A compilation of anonymized individual data is provided in Supplemental File S2^26^.

**Figure 2.**
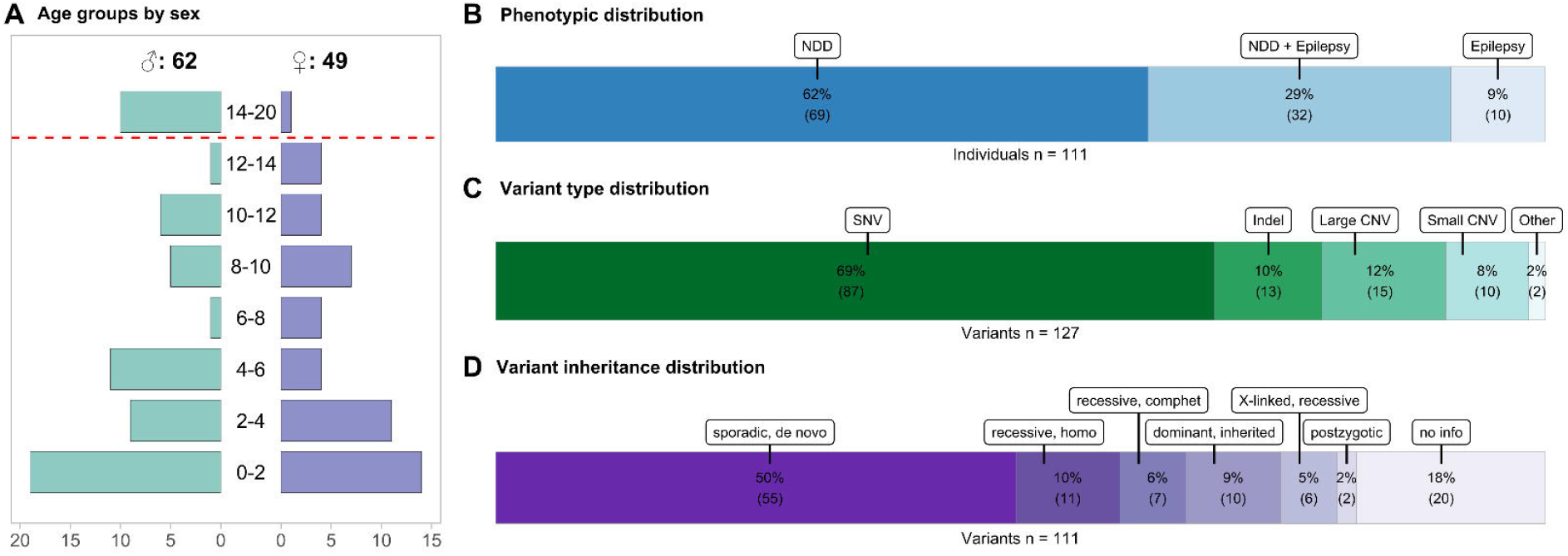
Individual and variant features. **(A)** Divergent plot representing the age distribution of individuals. The age determination refers to the date of final establishment of the human genetic diagnosis. The y-axis represents the age classes in years. The x-axis shows the number of associated individuals. **(B)** Stacked bar chart indicating the distribution of the phenotypes (epilepsy, NDD and the combination of both) in the 111 included individuals presented in percentages. **(C)** Stacked bar chart showing the distribution of all variant types found in the individuals including additional findings seen in no or low association with NDD or epilepsy presented in percentages (n=127). SNV = Single-nucleotide variant, Indel = Insertion/Deletion Polymorphism, Large CNV = Copy number variation whose alteration affects multiple genes, Small CNV = Copy number variation whose alteration affects a single gene and involves, for example, several exons **(D)** Stacked bar chart showing the inheritance patterns of those variants assigned to the respective phenotype of epilepsy or NDD (n=111). homo = homozygous, comphet = compound heterozygous.

### Genetic diagnostics

By study design all 111 individuals received their molecular diagnosis by NGS-based genetic diagnostics. Four individuals (3.6%) received an in-house custom-design panel targeting genes associated with epilepsy (“Epi-panel”, 122 genes). TruSight One v1 Panel (Illumina, Inc., San Diego, CA) was performed in 68 individuals (61.3%) which includes 4,811 genes associated with human disease. Whole exome sequencing (WES) targeting all coding genes was performed in 38 individuals (34.2%) using either a BGI Exome capture 59M kit (ten individuals; BGI, Shenzhen, China), SureSelect Human All Exon V6 (four individuals; Agilent Technologies, Santa Clara, CA, USA) or a TWIST Human Core Exome Kit (25 individuals; TWIST Bioscience, San Francisco, CA, USA) target design. For all included individuals, the genomic regions targeted by the respective enrichment design had an average coverage of ≥ 100 reads and ≥ 95% were covered by ≥ 10 reads.

### Variant characteristics

In 111 individuals, a total of 127 variants was reported through NGS-based diagnostics. The majority of 87 (68.5%) variants were single-nucleotide variants (SNV), with the remainder composed of 15 (11.8%) copy number variants affecting multiple genes (Large CNV), 13 (10.2%) insertion/deletion variants (indel), ten (7.9%) copy number variation whose alteration affects a single gene and involves several exons (Small CNV), and two complex events (1.6%) summarized as “other” (Figure 2C). These comprise an unbalanced translocation identified through coverage-based CNV analysis and subsequently confirmed by karyotyping and FISH and a deletion-insertion event in in *MECP2* (Variant ID: CNV017 and SNV008; Supplemental File S3^26^). Of these 127 variants, 111 variant combinations represented as causative for the phenotypes of NDD/epilepsy in the individuals, whereas the other genetic alterations were related to other concerns or were incidental findings. The origin of most NDD/epilepsy-related variants was recorded as sporadic and *de novo* in 55 cases (49.5%). The inheritance of eleven variants (9.9%) was recessive and homozygous, ten (9%) were dominant and inherited, another seven (6.3%) variants combinations represented as recessive compound heterozygous variants, six (5.4%) were X-linked recessive and two (1.8%) were postzygotic. For 20 variants (18%) no segregation was performed in the parents (Figure 2D). A detailed compilation of all identified variants is provided in Supplementary File S3^26^.

### Variants Reevaluation

Reevaluation resulted in downgrading of one variant previously reported as pathogenic and exclusion of one individual from subsequent analyses (Ind076; Supplemental File S3^26^) leaving 111 individuals for our final cohort. The variant is a microdeletion (CNV_013) identified in Ind076 which does not adequately explain the highly disabled individual’s phenotype in its severity. This individual deceased on the fourth day of life after uncontrollable seizures and severe asphyxia. The microdeletion identified, which includes *SIN3A* among others, is associated with milder disease cases.^30^ Because of the challenging assessability of this case due to the early death of the individual, we decided to exclude this case from our calculations, as we cannot fully clarify it. After reassessment of all variants formerly classified as (likely) pathogenic, 106 out of 111 phenotype-related variants remained in those assessment categories (Supplemental File S3^26^). Five variants had to be downgraded to VUS (Table 1). This can be explained inter alia by the change in recommendations regarding the PM2 criterion. A ClinGen^31^ recommendation to modify the strength of the PM2 criterion “Absent from controls, or at extremely low frequency if recessive, in Exome Sequencing Project, 1000Genomes Project, or Exome Aggregation Consortium”^29^ resulted in a downgrade from moderate to supportive. The PS2 criterion was betimes judged to be too strong in some but can only be assessed with strength moderate due to unspecific phenotype in most NDD entities and lack of data on the occurrence of this variant in other individuals. For further verification, RT-PCR would have been necessary for Ind069 to confirm the splice effect of the *de novo* variant c.4581+18A>G in *SCN1A*. Because these five variants were estimated to be very plausible with the phenotype despite their classification as VUS, we nevertheless considered these cases resolved (“hot VUS”) and retained these individuals in the cohort. Further investigation and more information on other individuals with the same variants will prospectively help to improve the assessment of these cases in the future.

**Table 1.** Information of the five individuals with “hot VUS” variants after reevaluation.

| Variant ID | Individual ID | Gene | Variant | Variant classification | Classification rules | Zygosity | Origin |
| --- | --- | --- | --- | --- | --- | --- | --- |
| SNV_002 | Ind002 | *TUBB2B* | c.611A>T, p.(Asn204Ile) | VUS | PM2_Supporting, PP2_Supporting, PP3_Supporting, PP4_Supporting | heterozygous | maternal |
| SNV_022 | Ind019 | *NPRL3* | c.434T>C, p.(Leu145Pro) | VUS | PS2_Moderate, PM2_Supporting, PP3_Supporting | heterozygous | *de novo* (confirmed) |
| SNV_027 | Ind025 | *DYNC1H1* | c.8945G>A, p.(Arg2982His) | VUS | PS2_Moderate, PM2_Supporting, PP2_Supporting, PP3_Supporting | heterozygous | *de novo* (confirmed) |
| SNV_067 | Ind069 | *SCN1A* | c.4581+18A>G, p.? | VUS | PS2_Moderate, PM2_Supporting, PP3_Supporting | heterozygous | *de novo* (confirmed) |
| SNV_073 | Ind077 | *NBEA* | c.4662+1G>C, p.Thr1519_Val1554del | VUS | PVS1_Moderate, PS2_Moderate, PM2_Supporting | heterozygous | *de novo* (confirmed) |

The re-evaluation of a *SMAD4* duplication initially reported as incidental finding in Ind012 (Supplemental File S3^26^, Supplemental Figure S1 and S2) showed that this was a retrotransposition event rather than a true genomic duplication of exons 2-3 and 5-12 as initially assumed. Alterations in *SMAD4* gene can cause the juvenile polyposis syndrome.^32^ The identification of the *SMAD4* variant as a retrotransposition removes the need for preventive examinations regarding polyps in the gastrointestinal system of the individual and its family. A correction report was submitted to the affected family based on the re-assessment in our study.

### Time periods in the diagnostic pathway

The total diagnostic time (first symptoms to final diagnosis) was split into subintervals (Interval 1-5; Figure 1B) to allow granular analysis of the diagnostic trajectory. The median duration between onset of symptoms associated with the underlying genetic disease and the final genetics report establishing the diagnosis was 34.1 months (range: 0.6 – 210.5; SD: 57.4). A median of 64.5 days (range: 8 – 395; SD: 60.8) passed between the initiation of NGS-based diagnostics leading to molecular diagnosis and the report of the molecular diagnosis.

We compared the length of subintervals in individuals with first symptoms before (n=56) and after (n=55) the availability of NGS-based diagnostics at UKL in April 2016 (Figure 3A). While Interval 4 (initiation of first genetic diagnostics to initiation of final genetic diagnostics) dominates in the total diagnostic duration for individuals with symptom onset before the establishment of NGS, Interval 1 (first symptoms to initial diagnostics) emerges prominently for individuals with first symptoms after April 2016. The duration of the total diagnostic trajectory (Interval 1-5) significantly (p < 0.001, Wilcox-Test) differed between individuals with first symptoms before and after 2016-04-01 (Figure 3B); the median interval length was 106.3 months (range: 21.7 – 201.5; SD: 50.2) for individuals with first symptoms before establishment of NGS, whereas it was shorter for individuals with first symptoms thereafter, with a median of 9.2 months (range: 0.6 – 43.3; SD: 12.1). This can be expected because recruitment of individuals in our study ended in August 2020. Therefore, the length of the total diagnostic time interval for individuals with first symptoms after the introduction of NGS is limited to a maximum of 4 years.

**Figure 3.**
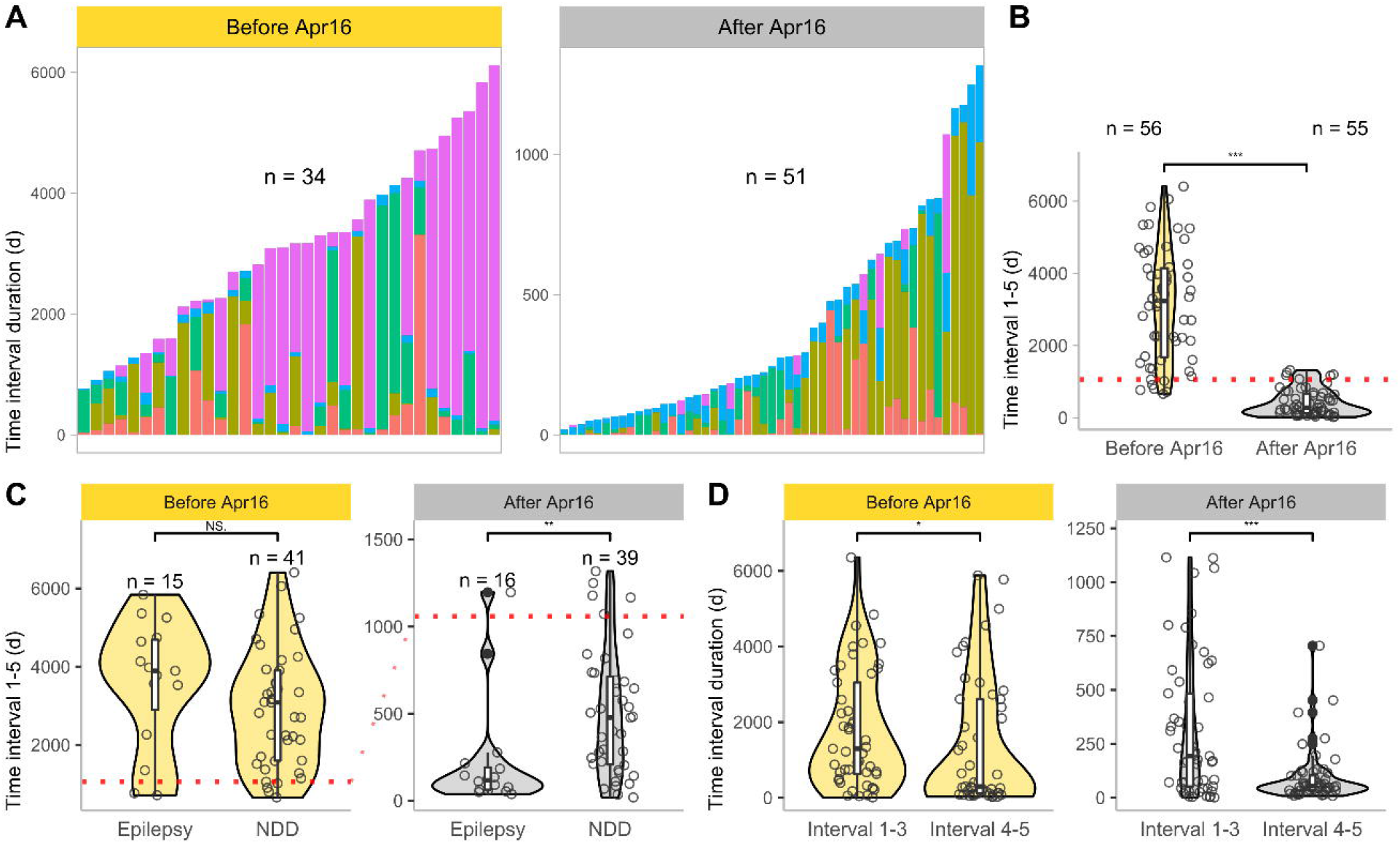
Impact of the establishment of NGS-based diagnostics on diagnostic time intervals. **(A)** Stacked bar plot showing the length of each diagnostic subinterval (Interval 1, 2, 3, 4 and 5) per individual, grouped by time of first symptom onset. The grouping was based on the establishment NGS-based human genetic diagnostics and labelled as before April 1, 2016 (Before Apr16) and after April 1, 2016 (After Apr16). Because of missing data in the medical histories of some individuals, we performed filtering for presentation purposes. Therefore, only individuals for which the duration of all diagnostic intervals could be calculated were included for this figure (n =84). The color selection of the diagnostic intervals is based on the colors of the timeline in Figure 1B. **(B)** Violin- and scatter-plot assembling the total diagnostic time interval (Interval 1 - 5, first symptoms to final human genetic diagnosis) grouped according to the time of onset of first symptoms. (p < 0.001, Wilcox-Test) **(C)** Violin- and scatter-plot assembling the total diagnostic time interval (Interval 1-5) grouped by phenotype according to the time of onset of first symptoms. Individuals with a combination of both phenotypes were assigned to the phenotype that occurred first in them, based on medical history research. (After Apr16: p ~ 0.002, Wilcox-Test) **(D)** Violin- and scatter-plot assembling Interval 1-3 (first symptoms to initiation of first genetic diagnostics) and Interval 4-5 (initiation of first genetic diagnostics to final diagnosis) grouped by the onset time of first symptoms. (After Apr16: p < 0.001, Wilcox-Test)

Dividing these subgroups based on their initial phenotype (NDD or epilepsy first), no significant differences between the phenotype groups emerged for individuals with initial symptoms before April 2016. Concerning individuals with first symptoms after 2016, those in the “epilepsy first” group received their molecular diagnosis significantly (p ~ 0.002, Wilcox-Test) faster (3.9 months; range: 0.13 – 36.53; SD: 10.6) than individuals in the “NDD first” group (15.7 months; range: 0.62 – 43.3; SD: 12) (Figure 3C).

To compare the proportion of the total diagnostic interval pertaining to pediatrics with that of clinical genetics, we attributed Interval 1-3 (pediatrics) and 4-5 (genetics) to them. While only a relatively weak significant difference (p ~ 0.03, Wilcox-Test) was seen between the pediatrics and genetics proportions for individuals with first symptoms before April 2016, a highly significant difference (p < 0.001, Wilcox-Test) was observed between Interval 1-3 and 4-5 for individuals with first symptoms after 2016. For these individuals, the time allocated to pediatrics (193 days; range: 1 – 1115; SD: 326.4) was significantly longer than that attributed to genetics (54 days; range: 8 – 705; SD: 123.2) (Figure 3D, Supplemental Figure S3). The duration of the total diagnostic pathway did not differ between individuals whose initial diagnosis took place at UKL and individuals whose initial diagnosis was performed at another medical institution in this cohort, in which all individuals received healthcare at UKL during the trajectory. (Supplemental Figure S4).

### Diagnostic procedures and costs

Before receiving a genetically confirmed diagnosis, the individuals in our cohort underwent various diagnostic procedures (Figure 4A, Supplemental File S2^26^). All individuals received at least one genetic examination, 101 individuals (91%) had at least one laboratory test other than metabolic testing and 86 individuals (77.5%) received at least one cMRI. Other medical imaging was performed at least once in 78 (70.3%) individuals, medical consults in 75 (67.6%), EEG in 74 (66.7%), metabolic diagnostics in 67 (60.4%), lumbar puncture in 26 (23.4%), electrophysiology in 26 (23.4%), functional tests in 14 (12.6%), ECG in 11 (9.9%). From the overall cohort 95 individuals (85.6%) were in inpatient care at UKL at least once (Supplemental Figure S5).

**Figure 4.**
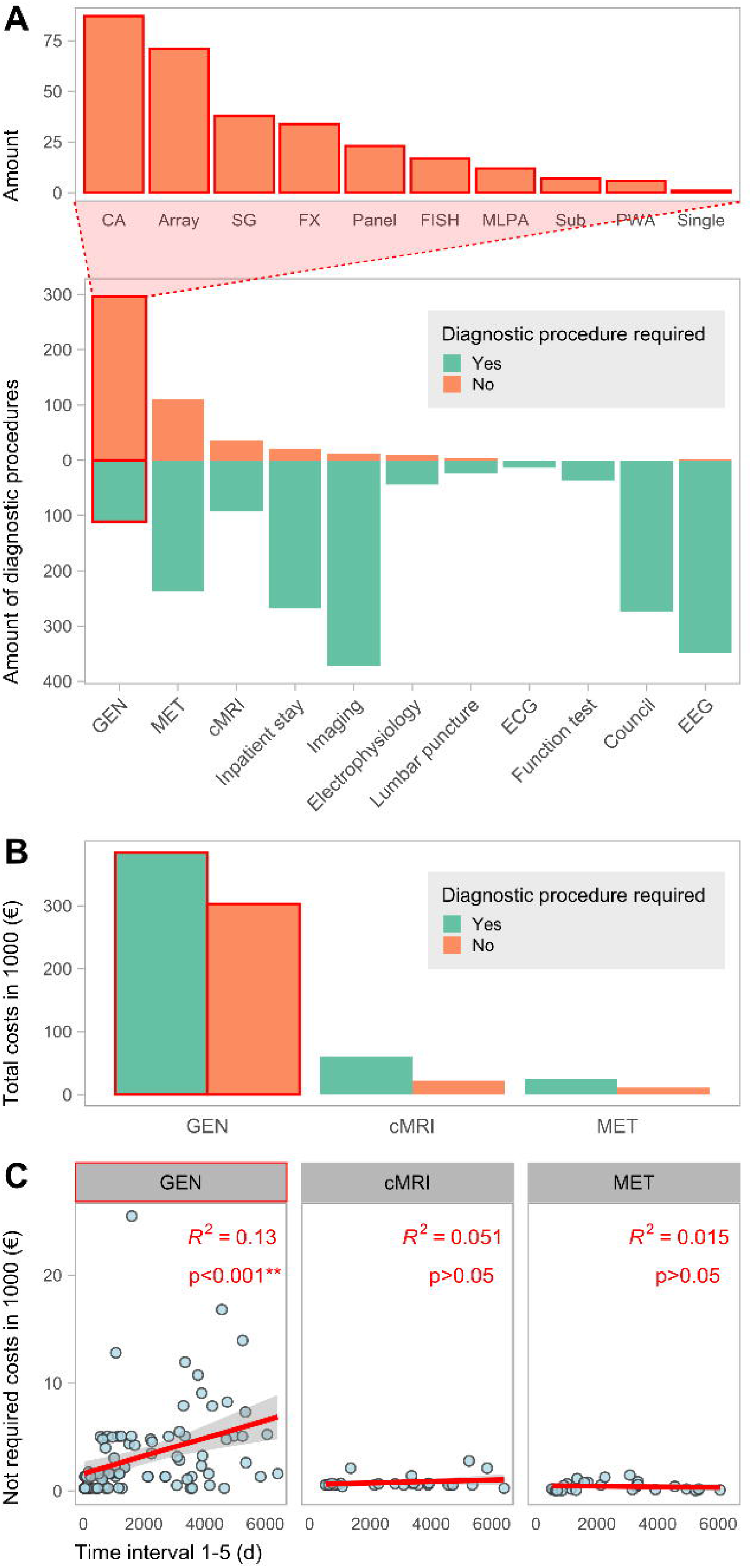
Required and non-required diagnostic costs. **(A)** Divergent plot illustrating the amount of required and not required diagnostics performed on the individuals. GEN = human genetic diagnostics, MET = metabolic laboratory testing, cMRI = cranial magnetic resonance imaging, Imaging = other medical imaging than cMRI, ECG = electrocardiogram, EEG = electroencephalogram. The bar chart above categorizes the non-required genetic diagnostics. CA = Chromosome analysis, SG = Single Gene analysis, FISH = Fluorescence *in situ* hybridization, MLPA = Multiplex ligation-dependent probe amplification, Sub = Analysis of Subtelomeres, PWA = Prader-Willi syndrome and Angelman syndrome diagnostics, Single = Single Exome Diagnostics. **(B)** Bar chart showing the respective costs grouped by top three diagnostic categories (with the highest amount of potential non-required diagnostics) and requirement. **(C)** Scatterplot showing the correlation of the respective unnecessary diagnostic costs with the length of the diagnostic odyssey (Interval 1-5), grouped by diagnostic category. The values for R^2^ and the p-values are given in the plot.

The diagnostic categories with the highest amounts of potentially non-required procedures are genetic diagnostics, metabolic diagnostics, and cMRI (Figure 4A). Of 407 genetic examinations performed overall, 296 (72.7%) were categorized as potentially dispensable if an exome-wide analysis would have been performed initially instead. The different types of dispensable genetic diagnostics are mainly constituted by chromosomal analyses, arrays, single gene testing, fragile X syndrome diagnostics and small custom gene panels (Figure 4A, top part). In total 129 cMRI examinations were performed in this cohort, of which 36 (27.9%) were categorized as not required. With one exception, all of those were performed under general anesthesia. Contrast agent was used in 18 (50%) of dispensable imaging procedures. Of 349 metabolic examinations, 111 (31.8%) were categorized as dispensable. Only a minority of procedures in the categories of laboratory tests other than metabolic tests, medical imaging except cMRI, electrophysiology, and EEG were classified as non-essential. All consults, ECG, functional tests, EEG examinations were classified as essential. We classified 21 (7.3%) of a total of 288 inpatient stays performed as potentially dispensable. Among these, non-required hospitalizations had a median of 2 inpatient overnight stays (range: 1 – 7; SD: 1.9) and were shorter, with a significant p-value (p ~ 0.016, Wilcox-Test), than hospitalizations classified as indispensable, with a median length of 4 nights (range: 0 – 731; SD: 46.7) (Supplemental Figure S6).

We calculated the costs for the three diagnostic categories with the largest number of non-required diagnostic procedures: genetic diagnostics, metabolic diagnostics, and cMRI. In our cohort, a total of 687,168.02€ was spent on genetic diagnostics. Thereof, 302,947.07€ (44.1%) are associated with dispensable examinations. Of the 82,589.20€ spent on cMRI, 21,903.37€ (26.5%) were considered not required if the final genetic diagnosis would have been known and considered. From 35,980.43€ issued for metabolic examinations, a portion of 10,987.05€ (30.5%) was classified as not required (Figure 4B). Thus, genetic examinations show the highest potential for cost savings in our cohort with 302,947.07€ (90.2%) out of 335,837.49€ in total potentially savable costs.

On average, the total potentially savable cost amounts to 3,025.56€ per individual in our study. This corresponds to an average of 2,729.25€ for genetic diagnostics, 197.33€ for cMRI examinations and 98.98€ for metabolic testing regarding potential cost savings.

The amount of summed dispensable cost per individual does not correlate with the length of the diagnostic trajectory (Interval 1-5) for metabolic diagnostics and cMRI. In genetic diagnostics, a moderate positive correlation (p < 0.001, Pearson correlation coefficient = 0.367, R^2^ = 0.13) is observed between the amount of the summed costs of genetic testing per individual and the length of time needed to obtain a diagnosis (Figure 4C).

## DISCUSSION

This retrospective cohort analysis emphasizes the importance of implementing ES as a first line test in the diagnostic pathway for individuals with NDD and/or epilepsy. NGS-based testing ended the hitherto inconclusive diagnostic pathway in all 111 individuals in our study. Therefore, the length of the diagnostic odyssey is significantly shorter in individuals with first symptoms onset after the availability of NGS than in individuals developing first symptoms before this cutoff time. It must be considered that our study included only individuals who received a final molecular diagnosis, and we can thus not assess individuals who had inconclusive NGS-based diagnostics. However, our design is a representative snapshot of the currently achievable diagnostic yield (~31-53%) using ES as a first-tier test^33^ in NDD/epilepsy.

Even before the broad availability of NGS in the clinical setting, genetic testing in this cohort was often initiated early by the treating pediatricians. However, the diagnostic odyssey was prolonged by the unavailability of genetic diagnostics covering the considerable heterogeneity observed in NDD and epilepsy (SysID database^34^ accessed on 2021-04-24 states 1,454 genes associated with NDD whereof 663 were associated with epilepsy). This caused a stepwise evaluation using karyotyping, microarrays and clinically suspected diagnoses. After the establishment of NGS, this phase receded, while the time interval before initiation of genetic diagnostics became critical for a fast diagnosis. Direct comparison of Interval 1-3 (pediatricians) and 4-5 (medical geneticists) before and after clinical availability of NGS demonstrates a remarkable reduction on the genetics side. This shows the accelerating impact of NGS-based methods on diagnostic time intervals. Thus, further potential for shortening the diagnostic process lies in the faster referral of individuals with NDD/ epilepsy to genetics testing.

Our data imply that individuals who first presented at other or smaller hospitals had no disadvantages in terms of elapsed time to molecular diagnosis. This achievement may be attributable to good cooperation between medical institutions or the pivotal and well-known role of our Center for Rare Diseases in our local area.

Individuals with epilepsy received a quicker diagnosis in our cohort than those with NDD after establishment of clinical NGS. Thus, the decrease in diagnostic duration is more apparent in individuals with epilepsy. This might be explained by the assumption that pediatricians may be more sensitive to the possible genetic background of epilepsy. The potentially more acute clinical presentation of epileptic seizures could also have led to a more rapid initiation of genetic diagnostics. Furthermore, the renowned focus on epilepsy research in genetics at UKL may have contributed to this. It would be desirable if this could be increasingly established for NDD entities without epilepsy in the future.

Stark et al.^35^ and Tan et al^36^ examined the cost-effectiveness of NGS and also considered potential cost savings by omitting other traditional diagnostic procedures. These studies did however not consider whether these diagnostic interventions might nevertheless be indispensable in individual cases. Soden et al.^27^ analyzed potentially dispensable medical costs through rapid ES in NDDs, but included diagnostics that would have been performed even if the genetic diagnosis was known in their cost savings model. Therefore, our careful reassessment of each individual case, represents the first study to examine the extent of diagnostics that could retrospectively be replaced by early implementation of ES based on accepted criteria for diagnostics of NDD and epilepsy in a tertiary Center for Rare Diseases. The greatest potential for cost savings concerns prior genetic diagnostics in our study. Of the genetic tests performed, 72.7% were classified as potentially dispensable representing 90.2% of total savable costs. A reversal from classical genetic diagnostics to an exome first approach^37^ would thus have resulted in reduced diagnostic costs in our cohort. Particularly because ES is also increasingly suitable for the detection of CNVs (in our cohort 26 CNVs, including small and complex alternations like translocations, were identified through NGS), which provides an alternative to chromosomal microarrays.^24,38–40^ In case of inconclusive results in first tier ES, genetic examinations could be extended accordingly to entities possibly not covered, such as translocations, repeat disorders (fragile X syndrome) or mitochondrial disorders.

The proportion of potentially dispensable metabolic (31.8%) and cMRI examinations (27.9%) is relevant in these categories. genetic testing can compete with, or even surpass, metabolic screening in terms of diagnostic yield.^39,41^ ES prior to the performance of these diagnostics should therefore explicitly be considered. Unlike other studies^42^ we did not classify all metabolic diagnostics as dispensable, because they are essential as confirmatory diagnostics and for decision making for therapies. Furthermore, metabolic testing currently still provides diagnostic results in critical situations faster than genetic testing. More rapid genetic results in the future could therefore replace additional metabolic examinations. Our data suggest that the potential for diagnostic savings through ES lies primarily in genetic diagnostics itself, which is an effect of the granulated and step wise approach performed historically. Except for metabolic and cMRI examinations, there was little to no potential for savings in other diagnostic categories.

A comparison of the savable costs in our cohort with the results of other studies is limited, because of differences in study design and cohort selection. Tan et al.^36^ and Stark et al.^35^ prospectively designed different diagnostic pathways in the course of ES in an undiagnosed cohort and compared their estimated costs. Based on this, cost savings per additional diagnosis (inflated and currency converted values: 6,237.14€ and 1,561.58€) were determined. Soden et al. provide the mean costs of prior negative testing in non-acute individuals (inflated and currency converted value: 17,785.18€). Chung et al^43^, Monroe et al.^42^ and Vrijenhoek et al.^22^ calculated the cost savings per individual resulting from avoidable medical examinations due to early ES implementation (inflated and currency converted values: 110.63€; 3,277.66€ and 5,090.39€). In some of these publications the costs of potentially unnecessary diagnostics even exceeded costs for ES. In our data, the cost of ES amounts to 3,461.45€ while the average cost savings by avoidable diagnostics is 3,025.56€ per individual. Thus, the average cost of potentially dispensable diagnostics almost reaches the cost level of an ES examination. The avoidable costs in this study are lower than in some previously publications. This may be an effect of our cautious evaluation of potentially dispensable diagnostics and our focus on solely direct diagnostic costs. Furthermore, previous studies report higher costs for individual parameters of metabolic diagnostics as well as for cMRI which could be explained by different pricing policies of health care systems in other countries.

Hospitalizations that we classified as potentially avoidable were shorter than indispensable inpatient stays. This can be explained by the solely diagnostic purpose of these not required hospitalizations, whereas required hospitalizations were mostly associated with complex therapies, emergency admissions, or a poor general condition of the individuals. While only 7.3% of inpatient stays (with a median of only 2 nights) were classified as dispensable, their potential omission could have been a major relief for affected families in individual cases.^44^ However, costs alone should not determine the course of action in the care of individuals affected by rare diseases. Early implementation of ES could not only reduce diagnostic costs and time, but also prevent exhausting or even risky procedures. In our study, more than a quarter of cMRI examinations were judged to be potentially dispensable and nearly all of these procedures were performed using anesthesia. The decision to put a child at risk^45^ for imaging with sedation and contrast agent could be influenced by the outcome of prioritized genetic testing since imaging results rarely lead to diagnosis in individuals with NDD.^46^ A quickly established molecular diagnosis can also affect therapy and further medical interventions^15,24,36,47^, have an impact on family planning^14,18^, and contribute to the psychological well-being of the parents.^16^

Moreover, our study demonstrates the importance of reanalyzing and reevaluating exome data.^14,48^ The reassessment of the *SMAD4* variant initially reported as incidental finding in in Ind012 is of decisive clinical importance for the affected family who now do not have to partake invasive colonoscopies anymore. In addition, our reevaluation revealed one individual in which the previously reported genetic diagnosis could not sufficiently explain the observed phenotype because the associated phenotype was too mild. The formal downgrading of five variants is expected and an effect of continuous development and stricter interpretation of the ACMG criteria for variant classification. Nevertheless, we considered the five cases as resolved by the plausibility of these “hot VUS”. Additional analyses in the future will facilitate a definitive assessment of these cases. Thus, reevaluating genetic findings in light of new guidelines and expanding genetic databases may yield new results.^49^

Limitations of this study include the relatively small cohort of 111 individuals, the lack of control group by focusing on solved cases only and the retrospective approach. Moreover, only a part of dispensable costs was determined, so the extent of these may potentially be higher. Some of the design choices in our study were imposed by the billing system and decentralized medical system in Germany.

Further studies on this topic should involve larger cohorts in a prospective setting. Collaboration among multiple rare disease centers and more complete collection of individuals’ medical data across the boundaries of single medical institutions would benefit for this research aim. However, individuals with rare NDD and/or epilepsy entities will surely benefit from continued development and research into rapid and effective diagnostic pathways. Therefore, close and informed collaboration between different medical specialties, such as pediatrics and human genetics, is essential. Both early consideration of a genetic differential diagnosis and quick performance of ES can contribute to reduce diagnostic time, costs and exhausting medical procedures and enable a sooner reproductive choice in the families.

## Data Availability

All data generated or analyzed during this study can be found in the online version of this article at the publisher's website.

## ACKNOWLEDGEMENTS

We thank all involved families for participating in this study. We appreciate the support of the Institute of Human Genetics team members Julia Herrmann, Uwe Herrmann, Dr. Julia Hentschel, Anja Heinze and Marcel Frömming. This manuscript is part of a doctoral thesis of Julia Klau.

## AUTHORS’ CONTRIBUTIONS

J.R.L. and S.B. conceived the initial study concept. J.K. and S.B. collected, curated and classified the clinical data. R.A.J. and M.R. provided the genetic data and performed submission to public databases, with updates performed by J.K. after reevaluation. J.K., H.O., M.R. and B.P. reevaluated all variants according to current standards. J.K., S.B. and B.P. cleaned, standardized and analyzed all clinical and genetic data. J.K. and B.P. created Figures 1, 2, 3 and 4 and all Supplementary materials. J.K., J.R.L. and B.P. wrote and edited the manuscript. All authors reviewed and commented on the final draft of the manuscript.

## WEB RESOURCES

AutoPVS1: http://autopvs1.genetics.bgi.com/

ClinGen CNV Interpretation Calculator: https://cnvcalc.clinicalgenome.org/cnvcalc/

ClinVar: https://www.ncbi.nlm.nih.gov/clinvar/

Decipher: https://www.deciphergenomics.org/

gnomAD: https://gnomad.broadinstitute.org/

Mutalyzer: https://mutalyzer.nl

Varsome: https://varsome.com/

## ABBREVIATIONS

ACMG: American College of Medical Genetics
cMRI: cranial magnetic resonance imaging
CNV: copy number variant
DRGs: Diagnosis Related Groups
ECG: electrocardiogram
EEG: electroencephalogram
ES: exome sequencing
GOÄ: Gebührenordnung für Ärzte
MLPA: multiplex ligation-dependent probe amplification
NDD: neurodevelopmental disorder
NGS: Next Generation Sequencing
qPCR: quantitative polymerase chain reaction
RT-PCR: reverse transcription polymerase chain reaction
SD: standard deviation
SNV: single-nucleotide variant
UKL: Leipzig University Medical Center
VUS: variants of uncertain significance

## SUPPLEMENTARY FILES

**File S1:** Supplementary notes with supplementary figures and tables.

**File S2:** Comprehensive tabular data describing relevant clinical cohort information (available^26^ on Zenodo).

**File S3:** Information on all variants identified in this cohort (available^26^ on Zenodo).

