## Supplementary material for "Exome first approach to reduce diagnostic costs and time – retrospective analysis of 111 individuals with rare neurodevelopmental disorders": File S1

### SUPPLEMENTARY FIGURES

#### Figure S1 Sequencing data of the *SMAD4* gene in Ind012 with retrotransposition-typical reads in exons 5 and 6

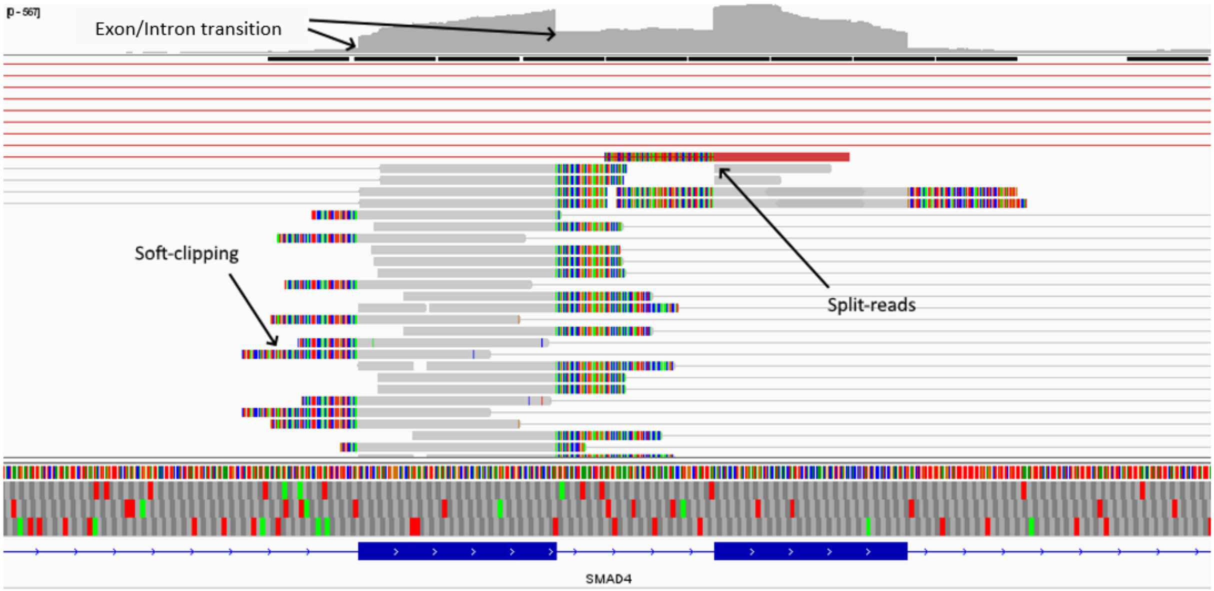


Sequencing data from individual Ind012 concerning questionable alterations in *SMAD4* gene. The variant previously reported as a duplication of exons 2,3 and 5-12 in the *SMAD4* gene is a retrotransposition and not a duplication of genomic DNA as originally thought. The sequence fragments ("reads") were visually inspected in the IGV browser. Here we observed that only the coding regions (including exon 4), but not the introns, show more of these reads. These unusual reads start in one exon and end in another ("split reads") or show a particular pattern in the mapping to the reference sequence with abrupt end of the sequence at the exon-intron transition ("soft-clipping"). This indicates the absence of introns and the presence of a spliced transcript at the DNA level. The additional reads still cause the bioinformatics algorithm to indicate a duplication. Taken together, this suggests the presence of a retrotransposon, which is characterized by the reverse transcription of processed mRNA with subsequent insertion into the genome and thus involves only the exons. These types of retrotransposons insert randomly into the genome, where they are very unlikely to be transcribed due to the lack of regulatory sequences. Furthermore, it is extremely unlikely to insert in the intact SMAD4 gene and disrupt its function here. In most cases, such retrotransposons are inherited, functionless and present in the non-coding region of the genome. In rare cases, it can insert into areas of other genes and thus interfere with their translation or transcription. For methodological reasons, the insertion point can not be determined by the method used.

**Figure S2 Position of the inconspicuous MLPA probe in intron 4 of the *SMAD4* gene**

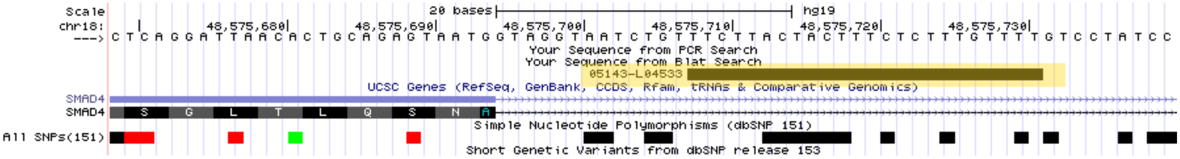


Moreover, the confirmation reported in our previous finding by a second method (MLPA Kit P158, company MRCHolland) and the observed absence of a duplication of exon 4 can be explained by the presence of a retrotransposition. Indeed, the probe for exon 4 in this kit ("05143-L04533", sequence: TTCTTACTACTT-TCTCTTTGTTTT) is located 24 base pairs after the exon-intron transition and is therefore not indicated as duplicated.

**Figure S3 Proportions of Interval 1-3 and Interval 4-5 in the total diagnostic interval (Interval 1-5)**

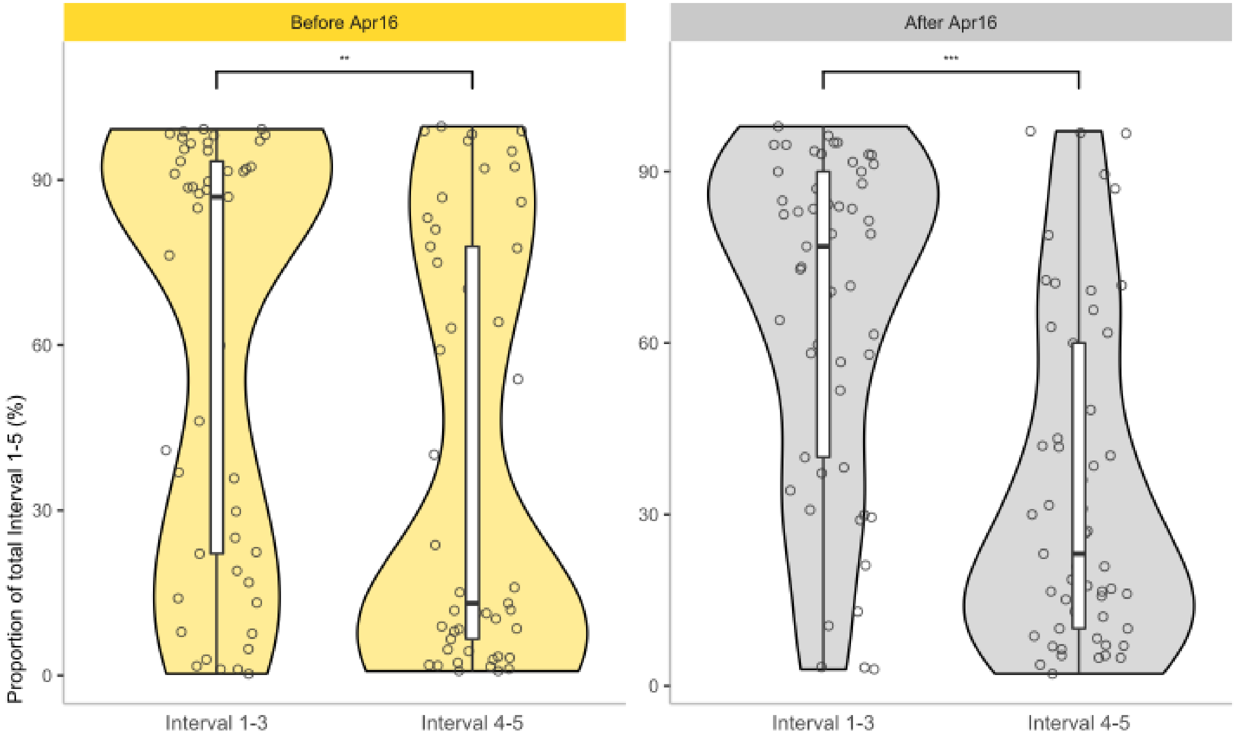


Violin- and scatter-plot showing the percentages of interval 1-3 and interval 4-5 within the total diagnostic interval grouped by the onset of first symptoms. The grouping was based on the establishment NGS-based human genetic diagnostics and labelled as before April 1, 2016 (Before Apr16) and after April 1, 2016 (After Apr16). Wilcoxon test was used for significant testing with significant results.

**Figure S4 Total diagnostic time interval grouped by the location of the initial diagnosis**

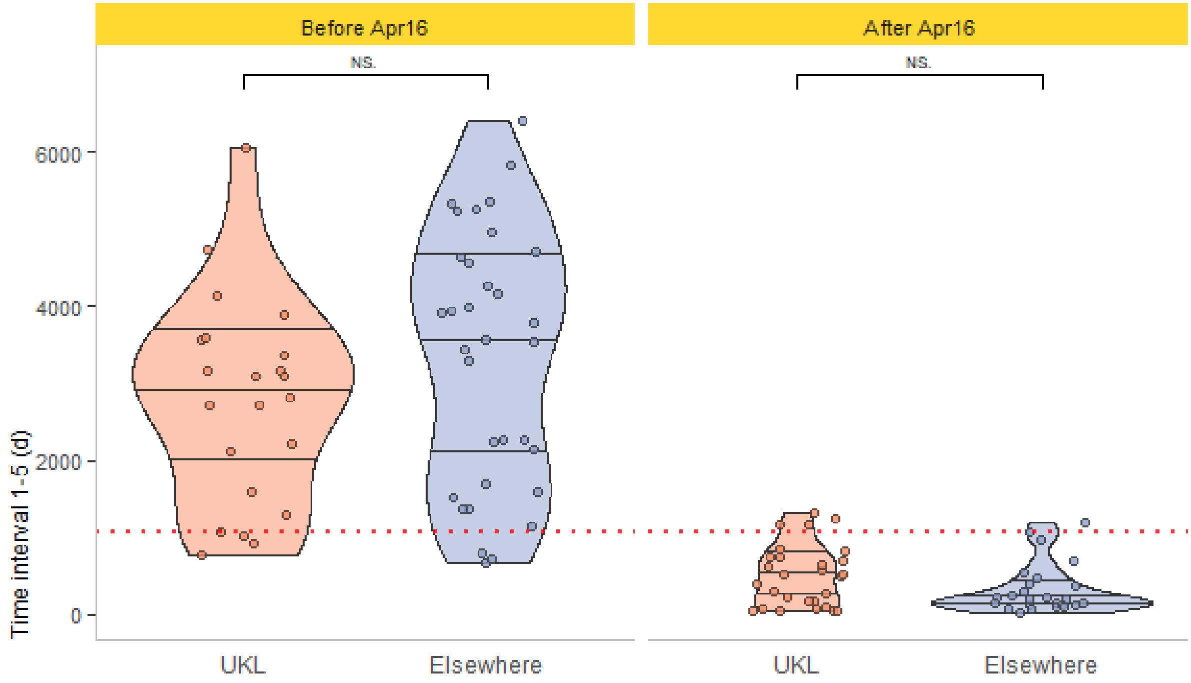


Violin- and scatter-plot assembling the total diagnostic time interval (Interval 1-5, first symptoms to final human genetic diagnosis) grouped according to the time of onset of first symptoms. The grouping was based on the establishment NGS-based human genetic diagnostics and labelled as before April 1, 2016 (Before Apr16) and after April 1, 2016 (After Apr16). The red dashed line marks the median of the total group. Wilcoxon test was used for significant testing with a non-significant result.

**Figure S5 Amount of performed diagnostic procedures**

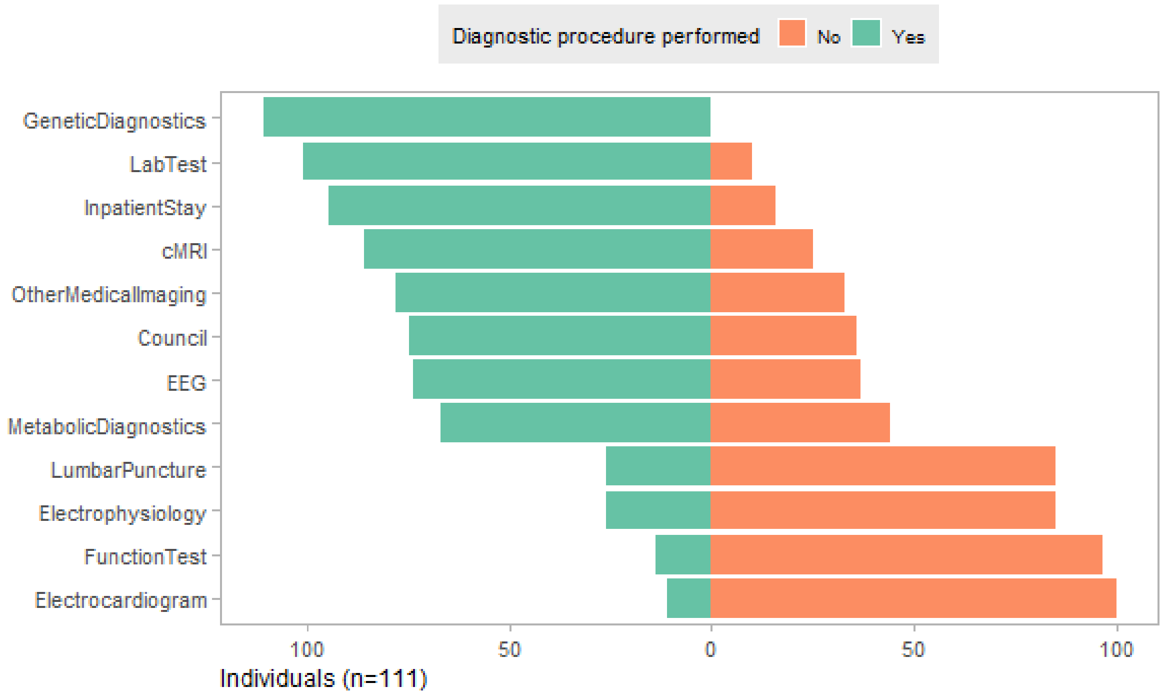


Divergent bar plot showing the amount of diagnostic measures performed and not performed. The left side in green indicates the proportion of the 111 individuals for whom the respective diagnostic procedure was performed at least once. The right side in green shows the remaining proportion of the cohort for whom the respective diagnostic procedure was never performed.

**Figure S6 Comparison of length of required and non-required inpatient stays**

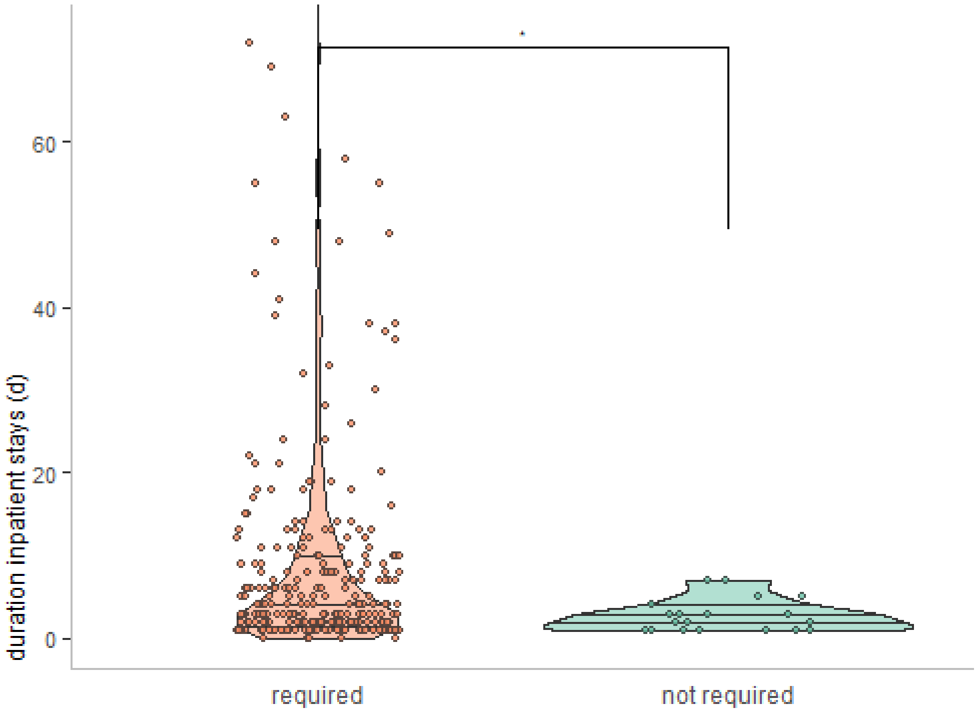


Violin- and scatter-plot assembling the length of hospital stays in days grouped according to our classification into required and potentially waivable inpatient stays. Wilcoxon test was used for significant testing with a weak significant result.

### SUPPLEMENTARY TABLES

**Table S1 Cost savings from other studies and their inflated values for 2021 in €**

| Study | Publication submission year | Cost type | Original costs reported | Inflated costs for 2021 | Inflated costs converted to € |
|---|---|---|---|---|---|
| Tan et al.[1] | 2017 | *cost savings per additional diagnosis* | US$6,838.00 | US$7,488.00 | €6,237.14 |
| Soden et al.[2] | 2014 | *average cost of prior negative tests in non-acute patients* | US$19,100.00 | US$21,352.00 | €17,785.18 |
| Chung et al.[3] | 2020 | *avoided healthcare costs* | HKD$1,005.53 | HKD$1034.26 | €110.63 |
| Monroe et al.[4] | 2015 | *average cost savings for genetic and metabolic investigations in diagnosed patients* | US$3,547.00 | US$3,935.00 | €3277.66 |
| Stark et al.[5] | 2016 | *cost savings per additional diagnosis* | US$1,702.00 | US$1,874.76 | €1561.58 |
| Vrijenhoek et al.[6] | 2017 | *avoided costs of traditional genotyping technologies, other diagnostic interventions, and all other laboratory investigations* | €4,896.00 | €5,090.39 | €5,090.39 |
